# Predicting Organ Rejections for Pediatric Heart Transplantations with a Combined Use of Transplant Registry Data and Electronic Health Records

**DOI:** 10.1101/2025.04.29.25326701

**Authors:** Balu Bhasuran, Xiaoyu Wang, Dipankar Gupta, Michael Killian, Zhe He

## Abstract

**Objective:** Pediatric heart transplantation is challenged by limited donor organ availability, prolonged waitlist times, and elevated risks of late acute rejection (LAR) and hospitalization. Current predictive models for post-transplant outcomes lack high accuracy due to reliance on registry data without integrating dynamic clinical and social factors. This study aimed to improve predictive performance and model interpretability by incorporating electronic health records (EHR), social determinants of health (SDoH), and United Network for Organ Sharing (UNOS) data.

**Materials and Methods:** We used EHR and UNOS data from 111 pediatric heart transplant patients (ages 0–18) at the University of Florida Health Shands Children’s Hospital to build predictive models for organ rejection at 1-, 3-, and 5-year intervals post-transplant. UNOS data includes pre- and post-transplant health and medical records, encompassing procedures, clinical evaluations, and post-transplant follow-up information, EHR data included evolving clinical parameters (e.g., comorbidities, medication adherence, and laboratory results), while SDoH encompassed socioeconomic status, living conditions, and healthcare access. Feature importance was assessed using Shapley Variable Importance Cloud (ShapleyVIC), which integrates Shapley Additive Explanations (SHAP) to provide robust, interpretable insights across nearly optimal models.

**Results:** Models integrating EHR, SDoH, and UNOS data outperformed those using UNOS data alone, with AUROC of 0.743 (0.607–0.879), 0.798 (0.725–0.871), and 0.760 (0.692–0.828). Key predictors of rejection included severe pre-transplant conditions (e.g., life support, prolonged waitlist times), elevated bilirubin and creatinine levels, and social factors (e.g., transportation barriers, BMI, insurance type).

**Discussion:** Findings reveal the importance of integrating clinical and social data to address multisystem dysfunction, disparities in healthcare access, and adherence challenges. ShapleyVIC enhanced model interpretability, providing actionable insights for improving post-transplant care.

**Conclusion:** Holistic, data-driven approaches that combine EHR, SDoH, and registry data significantly enhance predictive accuracy and interpretability, supporting improved long-term outcomes for pediatric heart transplant patients.

## Introduction

Advancements in pediatric solid organ transplantation have significantly improved survival rates, with heart transplant recipients achieving a five-year survival rate of approximately 81.5% between 2009 and 2013 [1]. However, these improvements are accompanied by persistent challenges, including high rates of late acute rejection (LAR) and recurrent hospitalizations [2–5], both of which can severely impact the quality of life for patients and their families [6–9]. Identifying patients at higher risk for adverse outcomes post-transplantation remains essential for optimizing care and delivering targeted interventions.

While data-driven modeling techniques have evolved, the clinical use of predictive models in pediatric transplantation remains limited [10–12]. Traditional approaches, such as general linear models and Cox regression, struggle to handle the complexity and heterogeneity of modern medical datasets [13–15]. In contrast, machine learning (ML) models excel at identifying hidden patterns within large datasets, offering more accurate predictions by capturing intricate interactions among clinical, demographic, and psychosocial factors. However, previous studies using ML models—such as random forests (RF) and adaptive boosting (AdaBoost)—have demonstrated only moderate success in predicting critical outcomes like rejection and mortality, with most relying on data from individual transplant centers, limiting their generalizability [16,17]. Additionally, deep learning (DL) models with national datasets have been underexplored. Our earlier work using national United Network for Organ Sharing (UNOS) data found that DL methods did not outperform models such as support vector machines (SVM), RF, or multilayer perceptron (MLP) in predicting post-transplant outcomes, including rejection and mortality at one, three, and five years [18].

In this follow-up study, we aim to enhance predictive performance and interpretability by integrating electronic health records (EHR) and social determinants of health (SDoH) with the UNOS dataset. Incorporating EHR data allows us to capture detailed clinical variables that evolve, such as comorbidities, medication adherence, and laboratory results, which are crucial in post-transplant outcomes. Additionally, integrating SDoH factors—such as socioeconomic status, education level, living condition, and access to healthcare—adds a critical dimension to understanding patient outcomes, as these non-clinical factors are known to influence post-transplant recovery and long-term health.

A key objective of this study is to address the limitations of prior research by leveraging a more comprehensive data landscape. To improve the interpretability of models with complex clinical data, we employ the Shapley Variable Importance Cloud (SHAP VIC) framework [19]. SHAP-VIC extends global interpretation beyond a single optimal model, pooling information across “good enough” models and visualizing the variability in variable importance, enabling more robust and practical assessments of clinical predictors. By leveraging SHAP-VIC, we ensure that the predictive models are both interpretable and reliable, providing actionable insights for clinical decision-making. Ultimately, by combining large-scale registry data with EHR and SDoH, we aim to develop personalized care strategies, reduce morbidity, and improve long-term outcomes for pediatric transplant recipients.

## Related Work

Previous research in pediatric heart transplantation has explored the UNOS registry and similar databases and applied machine learning models, such as random forests (RF) and adaptive boosting (AdaBoost), to predict post-transplant outcomes with varying success. Recent advancements in interpretable machine learning, including SHAP, have provided new frameworks for understanding model behavior, enabling more transparent and robust assessments of variable importance.

Miller et al. [12] utilized data from the UNOS Registry to predict mortality in pediatric heart transplantation using Random Forest (RF), Classification and Regression Trees (CART), and Artificial Neural Networks (ANN). Among these models, RF demonstrated the best performance, achieving AUROC of 0.72, 0.61, and 0.60 for 1-, 3-, and 5-year mortality predictions on the test set. However, the study reported low sensitivity across all models, attributing this to missing data, an outcome bias toward more common events, and imbalanced data. In a subsequent study, Miller et al. [20] analyzed outcomes in 8,349 pediatric heart transplant recipients from the UNOS database. They applied RF, XGBoost (Extreme Gradient Boosting), and L2-regularized logistic regression (LR) to predict all-cause mortality at 90 days and 1-year post-transplantation. Additionally, three survival analysis models—Random Survival Forest (RSF), Survival Gradient Boosting (SGB), and L2-regularized Cox regression (CR)— were employed to enhance predictions using time-to-event data. The models were evaluated using shuffled 10-fold cross-validation (CV) to ensure robustness and rolling CV (i.e., where a fixed-size window of data is moved sequentially through the dataset, with the model being trained on each window and evaluated on the subsequent data points outside the window) to simulate real-world, prospective performance. RF achieved the highest AUROC (0.893; 95% CI: 0.889–0.897) using shuffled CV, while XGBoost attained an AUROC of 0.657 (95% CI: 0.647– 0.667) with rolling CV. The study highlighted the performance decline because of temporal shifts in the data over time. Gupta et al. [11] investigated prolonged length of stay after pediatric heart transplantation, defined as hospitalizations exceeding 30 days, using the Pediatric Heart Transplant Society (PHTS) database. They developed models with stepwise logistic regression (SLR), gradient boosting (GB), and R), ultimately reporting a final model with an AUROC of 0.75 (95% CI: 0.72–0.78).

Killian et al. [17] explored the predictive performance of ML and DL models for post-transplant hospitalization across pediatric heart, kidney, and liver transplants using UNOS data. Traditional ML models such as RF, LR, MLP, and SVM were compared against a simple feedforward neural network (NN) model. For kidney transplants, the best models were LR (AUROC 0.62) for 1 year and RF (AUROC 0.55 and 0.60) for 3- and 5-year predictions. In the liver cohort, RF performed best for 1 year (AUROC 0.59), LR for 3 years (AUROC 0.63), and RF again for 5 years (AUROC 0.64). For heart transplant recipients, the top models were MLP (AUROC 0.72) at 1 year, and RF at both 3 and 5 years (AUROC 0.67 and 0.74, respectively). In another study, Killian et al. [18] used seven ML models and one DL model to predict both rejection and mortality at 1, 3, and 5 years post-heart transplantation. They employed Shapley additive explanation (SHAP) values to determine variable importance. RF emerged as the top-performing model in five out of six outcomes, covering 1-, 3-, and 5-year mortality as well as 1- and 3-year rejection, with AUROC values ranging from 0.664 to 0.763. AdaBoost performed best for 5-year rejection, achieving an AUROC of 0.705. Wisotzkey et al. [21] developed prediction models using the PHTS registry to assess 1-year graft loss-free survival. They implemented Cox regression, gradient boosting, axis-based random survival forests, and oblique random survival forests. SHAP values were employed for variable selection, importance ranking, and effect size estimation. The study further evaluated discrimination and calibration metrics to build the final prediction model, with RF showing the best performance, achieving a C-statistic of 0.74 (95% CI: 0.72–0.76).

## Methods

### UNOS Data (Transplant Center Data)

This study analyzed pediatric transplant data from the UNOS and EHR data, focusing on primary pediatric heart transplant (HT) recipients aged 0 to 18 years from the University of Florida Health Shands Children’s Hospital between 1988 and May 31, 2017 [22]. Exclusion criteria included patients who had undergone re-transplantation, cases with missing follow-up dates, patients without follow-up information within the prediction window, and those with unknown or missing values in their outcome variables. Transplant centers collected and reported data on pre-existing conditions, details of the transplant procedure, post-transplant complications, and long-term health outcomes. These data were organized into specific variables, which were then used as predictors and response variables.

### EHR and IDR Transplant Cohort Data Collection

We obtained linked EHR data with UNOS registry data for pediatric heart transplant patients. The EHR data contains demographics, encounters, diagnoses, lab test results, procedures, and medications. The UNOS dataset includes a range of individual, family, and medical factors aimed at predicting long-term health and transplant outcomes, specifically examining post-transplant hospitalization. UNOS collects both national and center-level data, tracking each patient from their listing for transplantation through annual follow-ups. Transplant centers are required to document pretransplant illness severity, details of the transplant procedure, postoperative events, complications, and health outcomes. Data for each patient is collected at three points: initial registration as a transplant candidate (TCR), registration at the time of transplant (TRR), and annual follow-up post-transplant (TRF). This structured data provides a comprehensive view of factors impacting pediatric transplant outcomes. EHR data and UNOS registry data were linked using MRN numbers and de-identified.

### Data Preprocessing

We performed a cross-check of the transplant date using CPT/ICPCS procedural codes in EHR and the transplant date from UNOS data and used the consent date as the index date. Then, we identified the first post-transplant organ rejection using ICD-9/ICD-10 diagnosis codes for rejection and used the date of the first rejection to determine whether a patient had organ rejection within 1-, 3-, 5-years post-transplant. We used the clinical features from EHRs within 6 months of pre- and post-transplant lab tests and 1-year post-transplant medications as well as features from the UNOS data (e.g., demographics of recipient and donor, recent graft status, primary diagnosis at listing, etc.). For each lab test, we calculated the median value for the 6-month pre- and post-transplant, respectively. Additionally, we have incorporated SDoH information from the clinical notes of the patients. After defining the patient cohort, we established the prediction outcomes and selected the observation and prediction windows. Next, we performed data normalization and imputation, followed by machine learning and model interpretation. The complete workflow for this study is illustrated in Figure 1.

**Figure 1:**
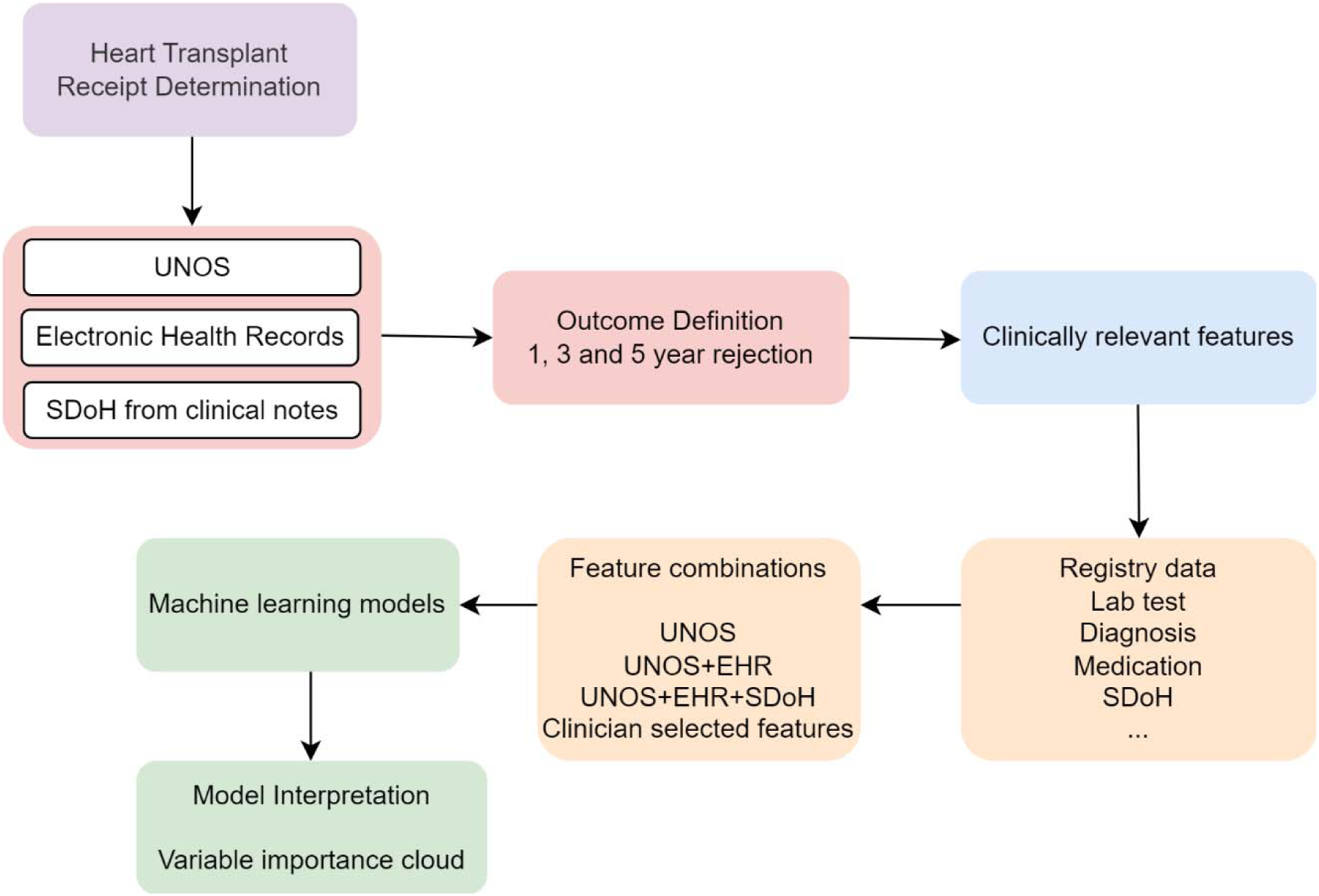
Study Pipeline

### SDoH Information Extraction from Clinical Notes

We leveraged a published open-source natural language processing package SODA (i.e., SOcial DeterminAnts) for extracting social determinants of health from clinical notes [23]. To develop SODA, seven transformer-based large language models (LLMs) (e.g., BERT_general, BERT_mimic, Roberta_general, Roberta_mimic, DeBERTa, GatorTron, and Longformer ) were fine-tuned and evaluated with an SDoH corpus consisting of 629 clinical notes of cancer patients with annotations of 13,193 SDoH concepts/attributes from 19 categories of SDoH, and another cross-disease validation corpus consisting of 200 notes from opioid use patients with 4,342 SDoH concepts/attributes. These 19 SDoH categories include: Abuse. Alcohol use, Drug use, Education, Ethnicity, Financial constraint, Gender, Language, Living condition, Living supply, Occupation/Employment, Physical activity, Race, SDoH ICD, Sexual activity, Social cohesion, Tobacco use, and Transportation. Among the evaluated transformer models, GatorTron achieved the best mean average strict and lenient F1 scores of 0.9122 and 0.9367 for SDoH concept extraction, and 0.9684 and 0.9593 for linking attributes to SDoH concepts. The cross-disease evaluation of concept extraction on the opioid test dataset also achieved satisfactory strict and lenient F1 scores of 0.8444 and 0.8760, making this tool a promising choice for our project.

Using 145,951 clinical notes from our cohort of 111 patients, SDoH information was extracted using the SODA package. This process generated both SDoH_standard_category (e.g., Gender, Living Condition, Education) and SDoH_raw_text (e.g., male, female; lives with his mother and maternal grandparents; lives with adoptive parents; 10th grade, 5th grade, high school). The extracted output was further filtered to exclude values such as “No,” “Not on file,” “Refused,” and “N/A.” For prediction, we included only SDoH categories present in 50% or more of the patients’ clinical notes. Details on SDoH presence across the cohort are depicted in Table 1. This table provides a breakdown of the distribution of different SDoH categories, highlighting their prevalence and completeness in our dataset.

**Table 1:** Social Determinants of Health (SDoH) categories, including the number and percentage of patients experiencing each standard SDoH category.

| SDoH Standard Category | Patients having SDoH | Percentage |
| --- | --- | --- |
| Living_supply | 111 | 100 |
| Marital_status | 111 | 100 |
| Education | 111 | 100 |
| Tobacco_use | 111 | 100 |
| Occupation | 111 | 100 |
| Living_Condition | 111 | 100 |
| Substance_use_status | 103 | 92.793 |
| Sex_act | 102 | 91.892 |
| Alcohol_use | 97 | 87.387 |
| Drug_use | 96 | 86.486 |
| Physical_act | 94 | 84.685 |
| Abuse | 89 | 80.18 |
| Sdoh_status | 85 | 76.577 |
| Financial_constrain | 84 | 75.676 |
| Transportation | 82 | 73.874 |
| Social_cohesion | 78 | 70.27 |
| Language | 77 | 69.369 |
| Employment_status | 26 | 23.423 |
| Sdoh_freq | 22 | 19.82 |
| Gender_status | 2 | 1.802 |
| Employment_location | 2 | 1.802 |
| Protection | 1 | 0.901 |

### Outcome definition

Figure 2 presents a comprehensive timeline for data collection, observation, and outcome prediction in this study, beginning at patient registration and extending through post-transplant periods. The timeline starts with the date of registration, marking when a patient is officially listed for a transplant. From this point, the observation window (pre-transplant) begins, encompassing data collected on various factors like UNOS data (medical history and demographics), phenotypes (observable traits related to health), medications (current prescriptions), labs (laboratory test results providing insights into health status), and SDoH (including lifestyle, economic stability, social support, and access to resources that may affect health outcomes). Approaching the date of transplantation, an additional observation window gathers more specific EHR data. This includes pre and post-transplant (6 months) lab test results, which offer critical information on health around the time of the procedure, and post-transplant (1 year) medications, which help manage recovery and prevent rejection. Following the transplant, outcome prediction windows assess health outcomes at 1-year, 3-year, and 5-year intervals. This structured approach enables a detailed understanding of the factors impacting short- and long-term outcomes, supporting predictive modeling across various timeframes to inform patient care and improve transplantation success.

**Figure 2:**
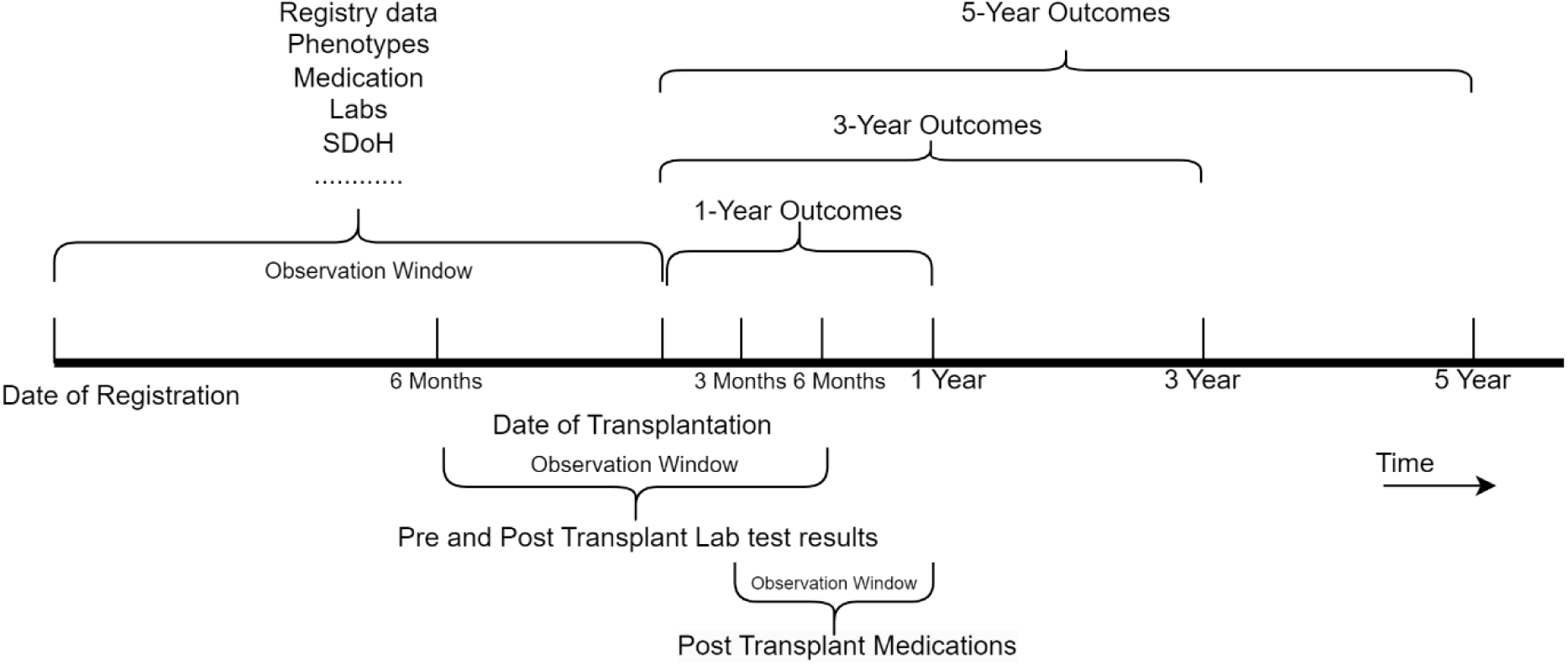
Observation window and outcome prediction windows.

### Feature Selection

This study aims to enhance predictive performance and interpretability by integrating EHR, SDoH, and UNOS data. This approach created a cohort with a large number of clinically relevant features. To evaluate the predictive accuracy of models, we incrementally incorporated features from various data sources. Initially, models were built using only features from the UNOS registry (previous study) [18]. In subsequent steps, additional datasets were combined in an additive manner, as described in Table 2. Each step builds on the previous dataset by adding more detailed data, ultimately resulting in a comprehensive dataset with features from diverse sources to optimize model performance and predictive accuracy. The selected features comprised both categorical and continuous numerical types. Categorical features were converted to numerical form for computational purposes. All continuous numerical variables were normalized to a range between 0 and 1. To address missing values, we applied multivariate imputation by chained equations. Following normalization and imputation, any variables showing collinearity with others were excluded. The final full study cohort consisted of 167 features.

**Table 2:**
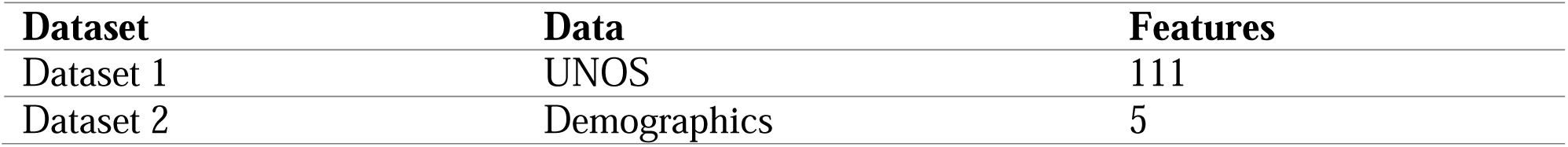
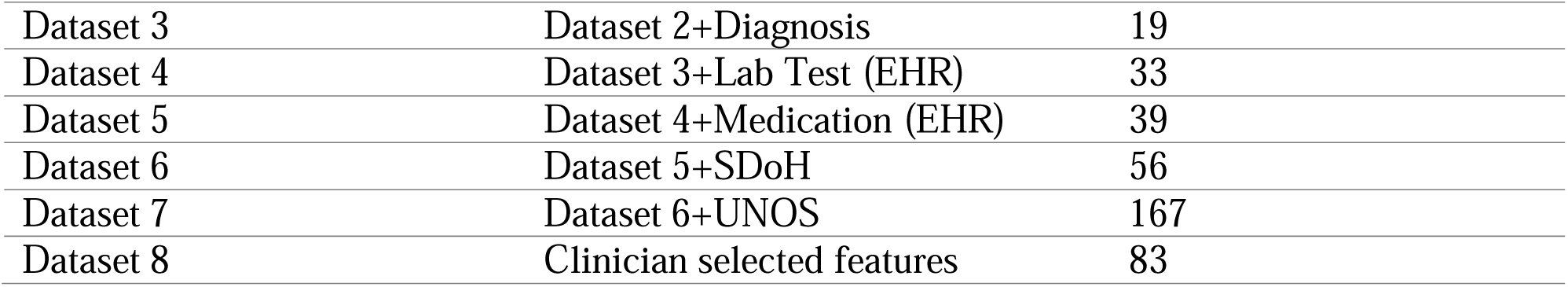
Data Sources and Features for Predictive Model Development.

| Dataset | Data | Features |
| --- | --- | --- |
| Dataset 1 | UNOS | 111 |
| Dataset 2 | Demographics | 5 |
| Dataset 3 | Dataset 2+Diagnosis | 19 |
| Dataset 4 | Dataset 3+Lab Test (EHR) | 33 |
| Dataset 5 | Dataset 4+Medication (EHR) | 39 |
| Dataset 6 | Dataset 5+SDoH | 56 |
| Dataset 7 | Dataset 6+UNOS | 167 |
| Dataset 8 | Clinician selected features | 83 |

### Machine Learning Models

In this study, seven machine learning (ML) models were tested: Logistic Regression, Random Forest, Support Vector Machine, AdaBoost, Gradient Boosting, XGBoost, and LightGBM. All models were implemented in Python using the scikit-learn package, with default settings applied. The evaluation metrics included Positive Predictive Value (PPV), Sensitivity, Specificity, F-score, Accuracy, and AUROC. A 10-fold cross-validation (CV) approach was used to evaluate each model. In each fold, 90% of the data was randomly selected for training, and the remaining 10% was used for testing. All evaluation metrics were calculated based on the 10-fold cross-validation results for each model.

### Model Interpretations

We used Shapley Variable Importance Cloud (ShapleyVIC), a novel approach to improve variable importance assessments in interpretable machine learning. Traditional methods focus on explaining only the final (optimal) model, potentially limiting insights in practical settings where “good enough” models are also valuable [24–26]. ShapleyVIC addresses this by extending Shapley-based variable importance across a set of “nearly optimal” models, providing a more robust measure of variable importance. This approach quantifies and visualizes uncertainty in variable importance, which helps to identify alternative models that balance accuracy and interpretability. ShapleyVIC integrates with Shapley Additive Explanations (SHAP) to assess variable contributions on both local and global levels, while accounting for variability across multiple models.

ShapleyVIC offers a comprehensive understanding of variable importance, reducing bias towards a single model and providing statistical inference capabilities. This method supports the development of interpretable models by highlighting variables that consistently impact predictions across well-performing models. The ShapleyVIC framework introduces a comprehensive approach to variable importance in interpretable machine learning by using a global importance measure, nearly optimal models, and a Variable Importance Cloud (VIC) [25]. The global importance measure, based on SAGE values, assesses each variable’s contribution across models that are “nearly optimal”—those within a small performance margin of the best-performing model (the Rashomon set). This approach goes beyond analyzing a single model by examining a range of high-performing models, offering insights into variables that consistently influence outcomes. VIC further enhances this by visualizing the variability in variable importance across the Rashomon set, highlighting how variable contributions can shift across different models. The average ShapleyVIC value of a variable represents its overall importance across nearly optimal models. The 95% prediction interval (PI) for a new model within the Rashomon set provides a statistical measure to evaluate and compare this importance. Since only positive values indicate significance, a variable’s overall importance is considered statistically significant when the lower bound of the 95% PI is positive. To illustrate these findings, a bar plot is used with error bars to display the average ShapleyVIC values alongside their 95% PIs. Additionally, a colored violin plot is included to visualize the distribution of model reliance (MR) and its relationship with model performance. This combined approach provides a robust, interpretable assessment of variable importance that minimizes bias toward any single model and has shown to be highly useful in clinical applications [19,27,28]. The current study reports the ShapleyVIC-based feature importance plots for explaining the model predictions for 1-year, 3-year, and 5-year rejection outcomes.

### Ethical Considerations

The study received approvals from the Institutional Review Board (IRB) at Florida State University under protocol number # STUDY00002444 and IRB at University of Florida under the protocol number #IRB202101911.

## Results

### Transplant Patient Cohort

Table 3 provides an overview of the characteristics of our pediatric heart transplant cohort consisting of 111 patients. The cohort comprises 63.06% males (n=70) and 36.93% females (n=41). In terms of racial distribution, 51.35% of the patients are White (n=57), 23.42% are African American and/or Black (n=26), 21.62% are Hispanic and/or Latino (n=24), 2.70% are Asian (n=3), and 0.90% are Multiracial (n=1). The majority of patients (67.56%, n=75) are in the 0–10 age group, while the remaining 32.43% (n=36) are aged 11–18 years. Congenital Heart Defect is the most common primary diagnosis (45.94%, n=51), followed by Dilated Cardiomyopathy (18.01%, n=20) and Hypertrophic Cardiomyopathy (4.50%, n=5). This distribution highlights a cohort primarily affected by severe pediatric cardiovascular conditions.

**Table 3:** Patient cohort characteristics.

| **Characteristics** | **Overall (N=111)** |
| --- | --- |
| **Gender, n(%)** |  |
| **Male** | 70 (63.06%) |
| **Female** | 41 (36.93%) |
| **Race, n(%)** |  |
| **White** | 57 (51.35%) |
| **African American and/or Black** | 26 (23.42%) |
| **Hispanic and/or Latino** | 24 (21.62%) |
| **Asian** | 3 (2.70%) |
| **Multiracial** | 1 (0.90%) |
| **Age, n(%)** |  |
| **[0-10]** | 75 (67.56%) |
| **[11-18]** | 36 (32.43%) |
| **Primary Diagnosis, n(%)** |  |
| **Congenital Heart Defect** | 51(45.94%) |
| **Dilated Cardiomyopathy** | 20(18.01%) |
| **Hypertrophic Cardiomyopathy** | 5(4.50%) |

### Model Performance

Figure 3 illustrates the mean receiver operating characteristic (ROC) curves generated from 10-fold cross-validation for various datasets from Table 2 and machine learning classifiers, evaluated over the three outcomes: (a) 1-year rejection, (b) 3-year rejection, and (c) 5-year rejection. Each subplot compares the performance of multiple classifiers, with the area under the curve (AUC) denoted for each dataset. The model performance of various machine learning models in these datasets is provided in Table 4. For 1-Year Rejection (Figure 3 a): Logistic Regression achieves the highest AUC of 0.76 on Dataset 1, while other classifiers such as Random Forest and Gradient Boosting exhibit varying performances across datasets, with AUCs ranging from 0.67 to 0.75. This suggests moderate predictive capabilities for 1-year outcomes, with Logistic Regression standing out. For 3-Year Rejection (Figure 3 b): The performance of classifiers improves slightly, with XGBoost achieving the highest AUC of 0.80 on Dataset 6. Random Forest, SVC, and AdaBoost show competitive AUCs, ranging from 0.65 to 0.79, indicating better long-term predictive accuracy compared to 1-year rejection. For 5-Year Rejection (Figure 3 c): XGBoost continues to perform well, with AUCs ranging from 0.76 to 0.79 across multiple datasets. Random Forest and SVC also demonstrate consistent performance, with AUCs ranging from 0.68 to 0.78, suggesting reliable predictions for long-term rejection outcomes. Overall, the figure highlights the variation in predictive performance across classifiers and datasets, with XGBoost showing robust performance for both 3-year and 5-year predictions. These findings emphasize the importance of model and dataset selection in optimizing predictive accuracy for transplant rejection at different time intervals.

**Figure 3:**
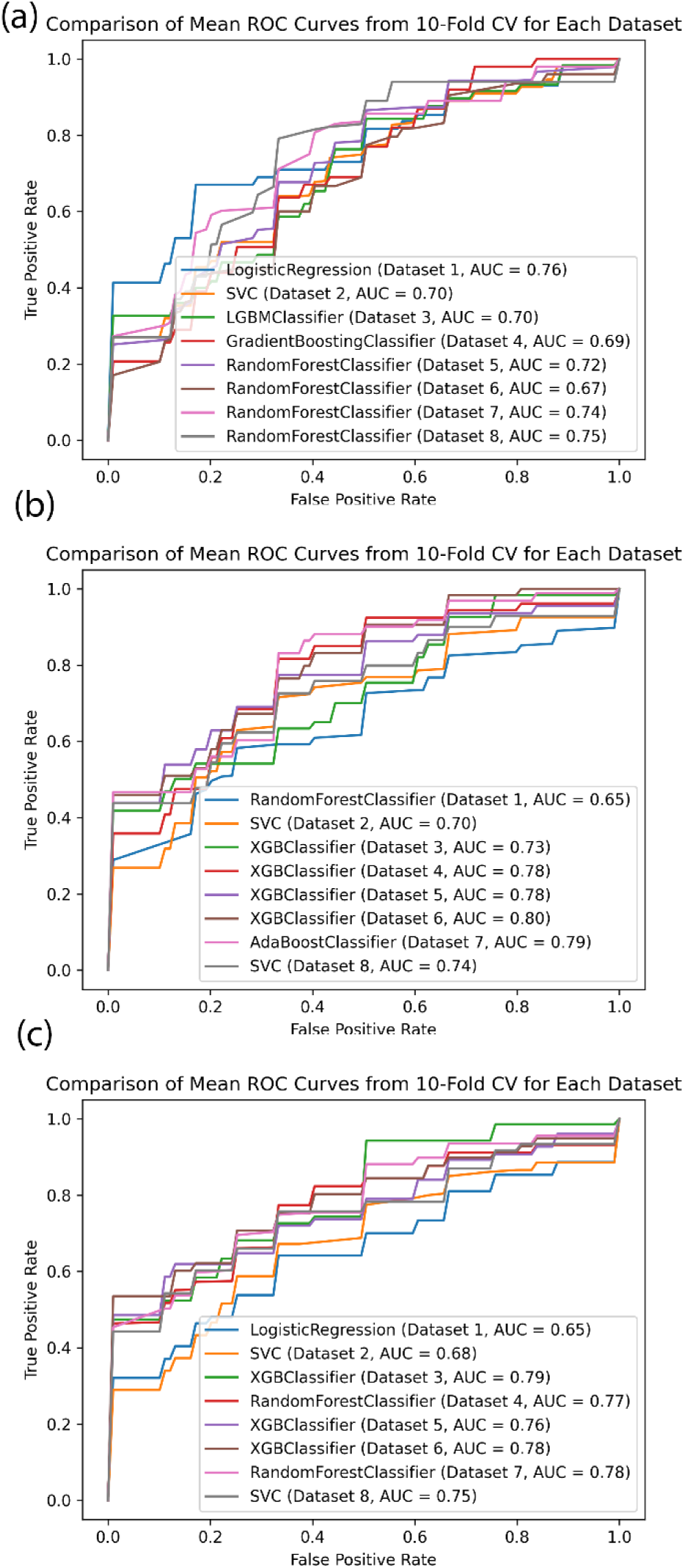
(a) Comparison of mean ROC curves from 10-fold cross-validation for Datasets 1-7 for 1-year rejection (b) 3-year rejection and (c) 5-year rejection

**Table 4:** Performance of different baseline prediction models for rejection.

| Outcome | Data | Model | Accuracy |
| --- | --- | --- | --- |
| **Rejection**<br>**At 1 Year** | UNOS | Logistic Regression | 0.711 |
|  | UNOS+ Demographics | XGBoost | 0.710 |
|  | UNOS+Demographics+ Phecode | Random Forest | 0.720 |
|  | UNOS+Demographics+ Phecode+ Lab Pre and Post | Random Forest | 0.702 |
|  | UNOS+Demographics+ Phecode+ Lab Pre and Post +Medications | Bagging Classifier | 0.711 |
|  | UNOS+Demographics+ Phecode+ Lab Pre and Post +Medications+SDoH | Gradient Boosting | 0.719 |
|  | UNOS+Demographics+ Phecode+ Lab Pre and Post +Medications+SDoH (Feature Selection) | Gradient Boosting | 0.720 |
| **Rejection**<br>**At 3 Year** | UNOS | Logistic Regression | 0.722 |
|  | UNOS+ Demographics | SVC | 0.722 |
|  | UNOS+Demographics+ Phecode | Random Forest | 0.730 |
|  | UNOS+Demographics+ Phecode+ Lab Pre and Post | Ada Boost | 0.720 |
|  | UNOS+Demographics+ Phecode+ Lab Pre and Post +Medications | Ada Boost | 0.738 |
|  | UNOS+Demographics+ Phecode+ Lab Pre and Post +Medications+SDoH | Ada Boost | 0.701 |
|  | UNOS+Demographics+ Phecode+ Lab Pre and Post +Medications+SDoH (Feature Selection) | Ada Boost | 0.748 |
| **Rejection**<br>**At 5 Year** | UNOS | Ada Boost | 0.766 |
|  | UNOS+ Demographics | SVC | 0.695 |
|  | UNOS+Demographics+ Phecode | LightGBM | 0.738 |
|  | UNOS+Demographics+ Phecode+ Lab Pre and Post | LightGBM | 0.719 |
|  | UNOS+Demographics+ Phecode+ Lab Pre and Post +Medications | LightGBM | 0.710 |
|  | UNOS+Demographics+ Phecode+ Lab Pre and Post +Medications+SDoH | LightGBM | 0.747 |
|  | UNOS+Demographics+ Phecode+ Lab Pre and Post +Medications+SDoH (Feature Selection) | XGBoost | 0.728 |

Table 5 compares the performance of machine learning models for predicting transplant rejection at 1 year, 3 years, and 5 years using evaluation metrics such as Positive Predictive Value (PPV), Sensitivity, Specificity, F-score, Accuracy, and AUROC, each with 95% confidence intervals (CI). For 1-year rejection, the Gradient Boosting Classifier achieved a PPV of 72.5% (95% CI: 0.53–0.911) and high specificity of 85.7% (95% CI: 0.760–0.953), but sensitivity was relatively low at 48% (95% CI: 0.34–0.618), resulting in moderate overall accuracy (71.9%) and AUROC (0.7429). For 3-year rejection, the AdaBoost Classifier showed improved sensitivity (66.7%, 95% CI: 0.503–0.830), balanced specificity (75%, 95% CI: 0.609–0.891), and higher AUROC (0.798, 95% CI: 0.725–0.871), indicating strong discriminatory power. The 5-year rejection model, based on LightGBM, demonstrated the best overall performance, with a PPV of 78.52% (95% CI: 0.713–0.858), sensitivity of 76.9% (95% CI: 0.684–0.854), F-score of 0.768 (95% CI: 0.727–0.809), and accuracy of 74.7% (95% CI: 0.704–0.790), along with an AUROC of 0.760 (95% CI: 0.692–0.828). These findings highlight the increasing predictive performance of models over longer time horizons, with LightGBM emerging as the most reliable model for long-term rejection prediction due to its robust balance of precision, sensitivity, and overall accuracy.

**Table 5:** Performance of the best prediction models for rejection in 1, 3, and 5 years.

| Outcome | Model | PPV<br>(95% CI) | Sensitivity<br>(95% CI) | Specificity<br>(95% CI) | F-score<br>(95% CI) | Accuracy<br>(95% CI) | AUROC<br>(95% CI) |
| --- | --- | --- | --- | --- | --- | --- | --- |
| Rejection<br>At 1 Year | Gradient<br>Boosting | 0.725(0.53-0.911) | 0.480(0.34-0.618) | 0.857(0.76-0.953) | 0.546(0.418-0.675) | 0.719(0.640-0.798) | 0.743(0.607-0.879) |
| Rejection<br>At 3Year | Ada<br>Boost | 0.755(0.636-0.874) | 0.667<br>(0.503-0.83) | 0.750(0.609-0.891) | 0.681<br>(0.573-0.788) | 0.701(0.608<br>- 0.795) | 0.798(0.725-0.871) |
| Rejection<br>At 5 Year | Light<br>GBM | 0.7852(0.713-0.858) | 0.769<br>(0.684-0.854) | 0.720(0.599-0.841) | 0.768<br>(0.727-0.809) | 0.747(0.704-0.79) | 0.760(0.692-0.828) |

Figures 4–6 illustrate the feature importance for predicting 1-year, 3-year, and 5-year rejection outcomes using ShapleyVIC. Figures 4a, 5a, and 6a depict the average ShapleyVIC values (with 95% prediction intervals, PIs) across 183 nearly optimal models (see SFigure 1).

**Figure 4.**
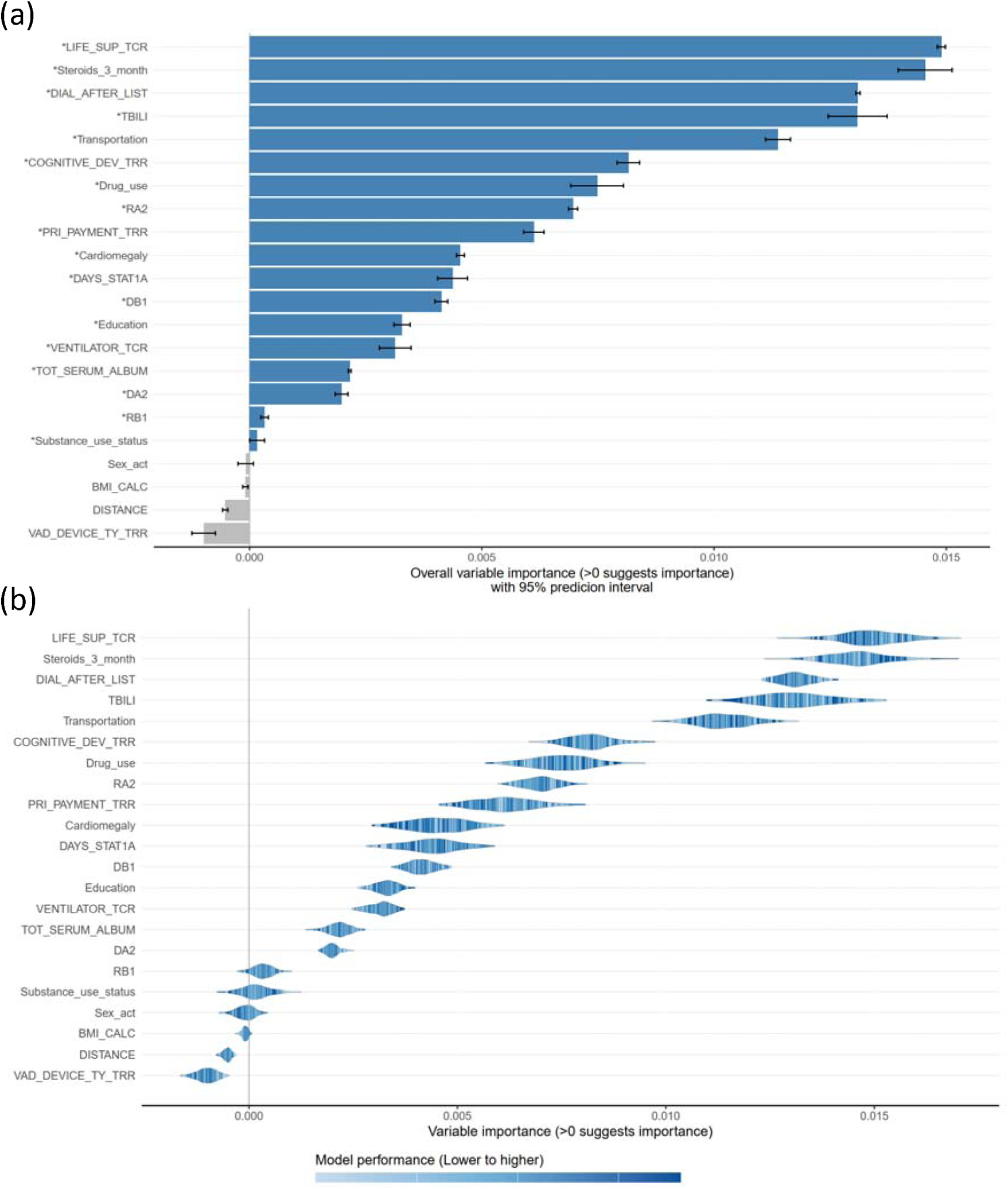
Visual summary of 1-year rejection prediction results using the ShapleyVIC framework. (A) Bar plot of ShapleyVIC values illustrating feature importance, accounting for variability across 183 nearly optimal models. The error bars represent 95% prediction intervals, indicating the range of variability in variable importance. (B) Violin plots showing the distribution of variable importance (shape and spread) across the nearly optimal models, with color gradients representing model performance (darker blue indicates better performance). The presence of dark blue strips towards the right end of a violin indicates that well-performing models relied heavily on the corresponding variable. This complementary visualization provides insights into both the variability and performance impact of each variable.

**Figure 5.**
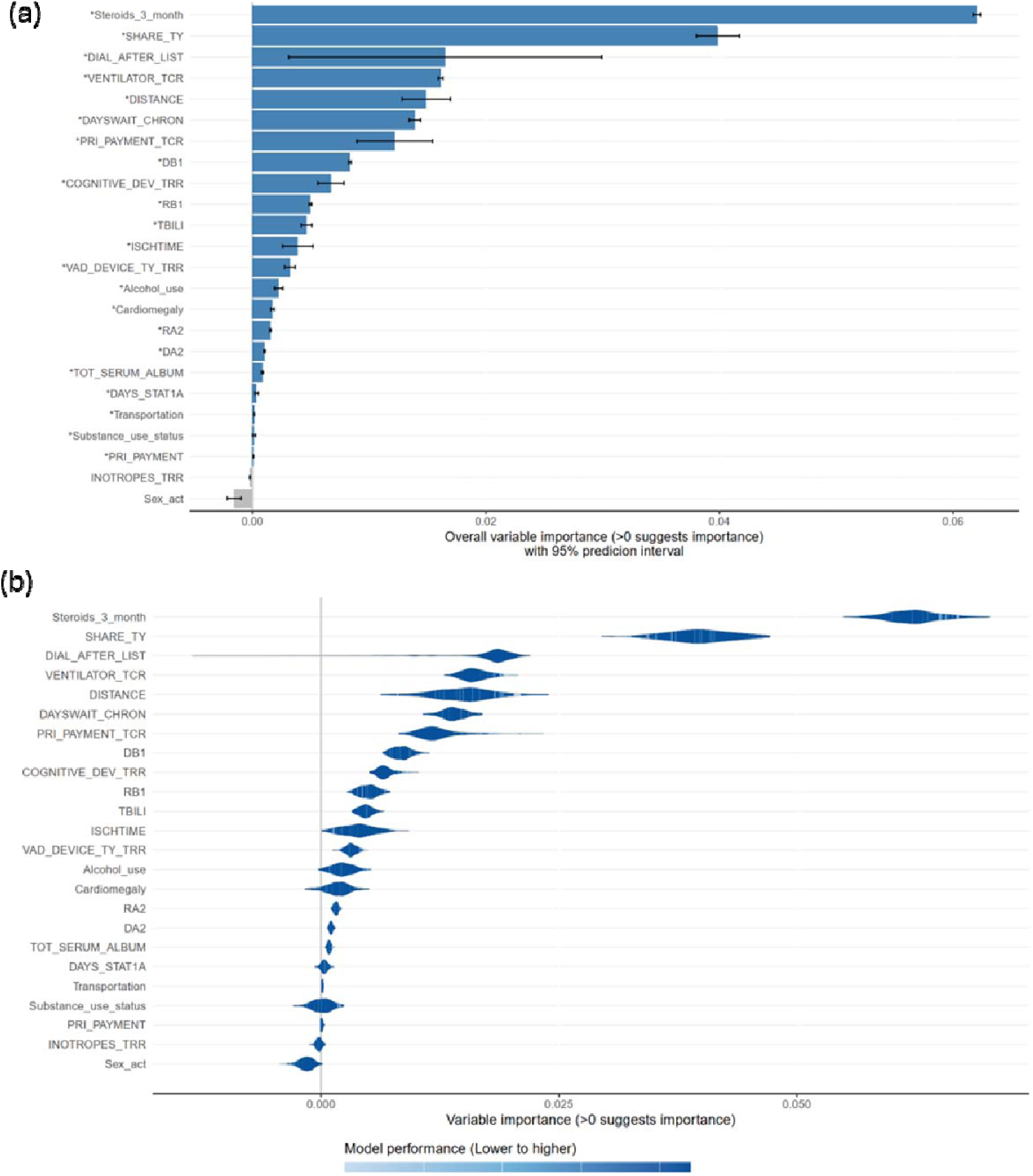
Visual summary of 3-year rejection prediction results using the ShapleyVIC framework. (A) Bar plot of ShapleyVIC values illustrating feature importance, accounting for variability across 183 nearly optimal models. The error bars represent 95% prediction intervals, indicating the range of variability in variable importance. (B) Violin plots showing the distribution of variable importance (shape and spread) across the nearly optimal models, with color gradients representing model performance (darker blue indicates better performance). The presence of dark blue strips towards the right end of a violin indicates that well-performing models relied heavily on the corresponding variable. This complementary visualization provide insights into both the variability and performance impact of each variable.

**Figure 6.**
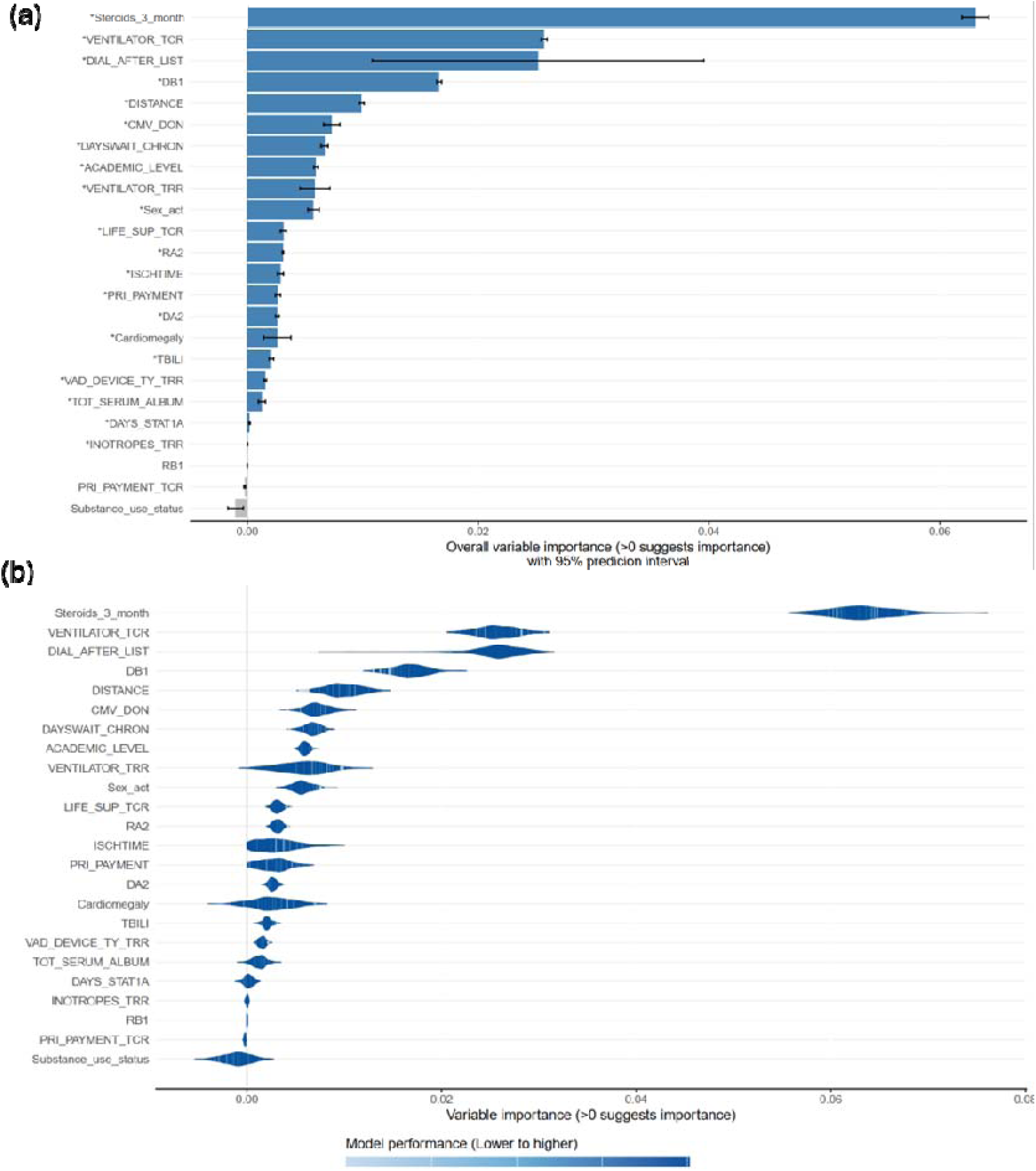
Visual summary of 5-year rejection prediction results using the ShapleyVIC framework. (A) Bar plot of ShapleyVIC values illustrating feature importance, accounting for variability across 183 nearly optimal models. The error bars represent 95% prediction intervals, indicating the range of variability in variable importance. (B) Violin plots showing the distribution of variable importance (shape and spread) across the nearly optimal models, with color gradients representing model performance (darker blue indicates better performance). The presence of dark blue strips towards the right end of a violin indicates that well-performing models relied heavily on the corresponding variable. This complementary visualization provides insights into both the variability and performance impact of each variable.

SFigure 1 illustrates the logistic losses of nearly optimal logistic regression models—defined as those with a loss within 5% (i.e., between 1 and 1.05 times) of the optimal model’s minimum loss. Figures 4b, 5b, and 6b visualize the variability in variable importance across models using violin plots, which show the relationship between model reliance (MR) on each variable and model performance. In the violin plots, the horizontal spread represents the range of MR for a variable, divided into equal-width slices. The height of each slice indicates the proportion of models within that MR interval, while the color reflects the average performance (in terms of empirical loss). Slices near the ends of the violins, which may lack models, are merged with adjacent slices closer to the center.

**SFigure 1:**
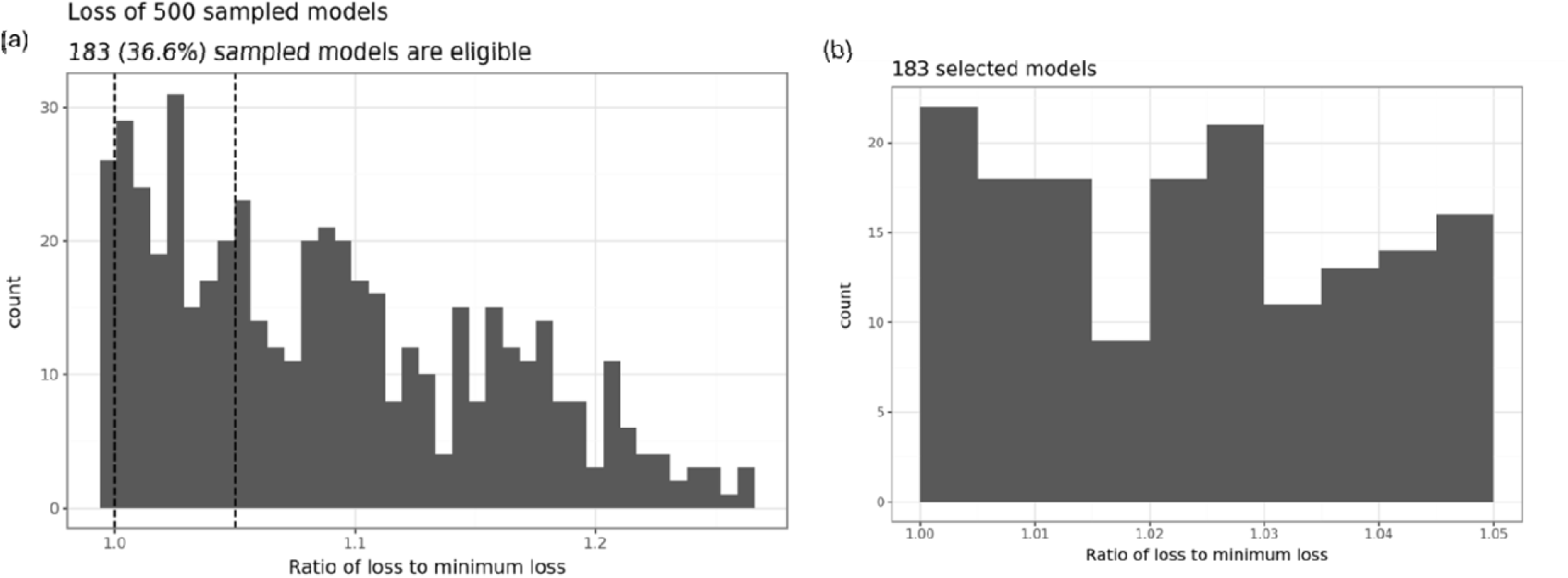
Loss of sampled nearly optimal logistic regression models with logistic loss less than times the minimum loss (i.e., logistic loss of the optimal model). The default value is 5% (1-1.05 ratio of loss to minimum loss).

In Figure 4a, features such as ‘LIFE_SUP_TCR’ (Candidate Life Support at Registration), ‘DIAL_AFTER_LIST’ (Dialysis Occurring Between Listing And Transplant), and ‘TBILÌ(Most Recent Serum Total Bilirubin at Transplant) show high average importance with PIs fully above zero, confirming their statistically significant contributions to predicting 1-year rejection. In contrast, features such as ‘DISTANCÈ (Distance From Donor Hosp To Transplant Center), ‘VAD_DEVICE_TY_TRR’(Ventricular Assist Device at Transplant Recipient Registration), and ‘Sex_act’ exhibit PIs near or below zero, indicating low or negligible importance. The variability in PIs, particularly for ‘DIAL_AFTER_LIST’, reflects differences in importance across models, suggesting variability in model reliance. Figure 4b demonstrates these distributions through violin plots, where features such as ‘LIFE_SUP_TCR’ show broad and uniform reliance across models, evidenced by a wide violin with consistent color, indicating robust importance. Conversely, features like ‘DISTANCÈ and ‘VAD_DEVICE_TY_TRR’ have narrow distributions near zero, reflecting limited relevance in most models. The color gradient highlights the relationship between reliance and performance; well-performing models tend to rely less on low-importance features like ‘DISTANCÈ.

In Figure 5a, features such as ‘Steroids_3_month’, ‘DIAL_AFTER_LIST’, and ‘VENTILATOR_TCR’(Patient On Life Support - Ventilator at Registration) emerge as the most influential for predicting 3-year rejection, with high ShapleyVIC values and PIs fully above zero, signifying statistical significance. In contrast, features like ‘PRI_PAYMENT_TCR’(Recipient Primary Payment Source at Transplant) and ‘Substance_use_status’ show minimal ShapleyVIC values and PIs crossing zero, indicating their limited relevance. Variability in PIs for intermediate features, such as ‘DISTANCÈ, reflects differences in their impact across models, guiding potential inclusion or exclusion in model refinement. Figure 5b reinforces these observations, with features like ‘Steroids_3_month’ demonstrating broad reliance distributions and consistent importance across models. In contrast, features such as ‘Substance_use_status’ have narrow distributions near zero, underscoring their negligible contribution. The color gradients within the violins reveal a clear trend: well-performing models depend less on low-importance features, such as ‘DISTANCÈ.

Similarly, Figure 6a highlights the importance of features such as ‘Steroids_3_month’, ‘DIAL_AFTER_LIST’, and ‘VENTILATOR_TCR’ for 5-year rejection predictions, with high ShapleyVIC values and PIs fully above zero, confirming their significant contributions across nearly optimal models. Conversely, features such as ‘Substance_use_status’ and ‘PRI_PAYMENT_TCR’ show minimal or negative ShapleyVIC values, with PIs crossing zero, indicating limited influence. Variability in PIs for features like ‘DISTANCÈ reflects their inconsistent importance, suggesting their limited reliability for prediction tasks. Figure 6b shows violin plots where features like ‘Steroids_3_month’ exhibit broad distributions with consistently high reliance, affirming their critical role in predictions. In contrast, features such as ‘Substance_use_status’ and ‘PRI_PAYMENT_TCR’ show narrow distributions near zero, reinforcing their limited relevance. The color variations within the violins show that higher reliance on key features, such as ‘Steroids_3_month’, aligns with better model performance, validating their predictive importance.

The error bars in Figures 4a, 5a, and 6a provide additional insights into the variability and statistical significance of variable importance across models. Variables such as ‘Steroids_3_month’, ‘DIAL_AFTER_LIST’, and ‘LIFE_SUP_TCR’ exhibit short error bars, indicating consistent importance and low variability across models, making them reliable predictors. In contrast, features like ‘DISTANCÈ and ‘VENTILATOR_TRR’ display longer error bars, highlighting substantial variability in their contributions, depending on the model configuration. Error bars crossing zero, as seen in features like ‘Substance_use_status’ and ‘PRI_PAYMENT_TCR’, indicate negligible or non-significant importance, suggesting these features are context-dependent or interact with other features inconsistently. Overall, the error bars and violin plots emphasize the importance of accounting for variability and uncertainty in assessing variable importance, ensuring robust, interpretable machine learning models for predicting rejection outcomes. The frequency of ranking of each variable for the 1-, 3- and 5-year rejection based on pairwise comparison of model reliance is depicted in Figures 7–9. The figure illustrates the ranking distribution of features across the optimal model set. Each bar represents how frequently a specific ranking is assigned to a feature among these models. Features with a tall bar on the left are consistently ranked highly across a large number of models, indicating their importance. Conversely, features with taller bars on the right are ranked lower, suggesting less importance in the models. If a feature has only one bar in its panel, it signifies that the feature’s rank remains constant across all the optimal models.

**Figure 7.**
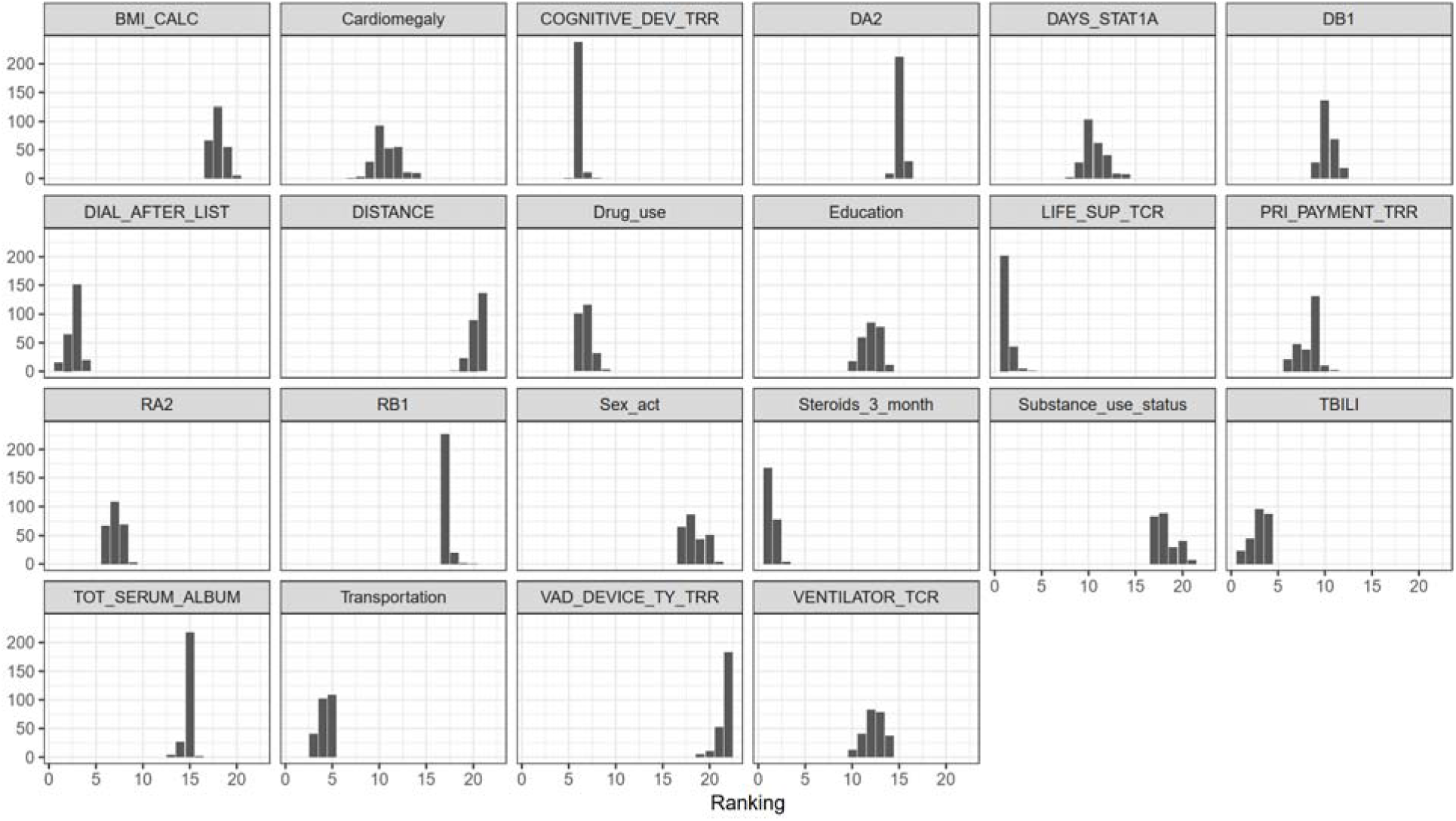
Frequency of ranking of each variable for the 1-year rejection based on pairwise comparison of model reliance. Variables are arranged by average ShapleyVIC values.

**Figure 8.**
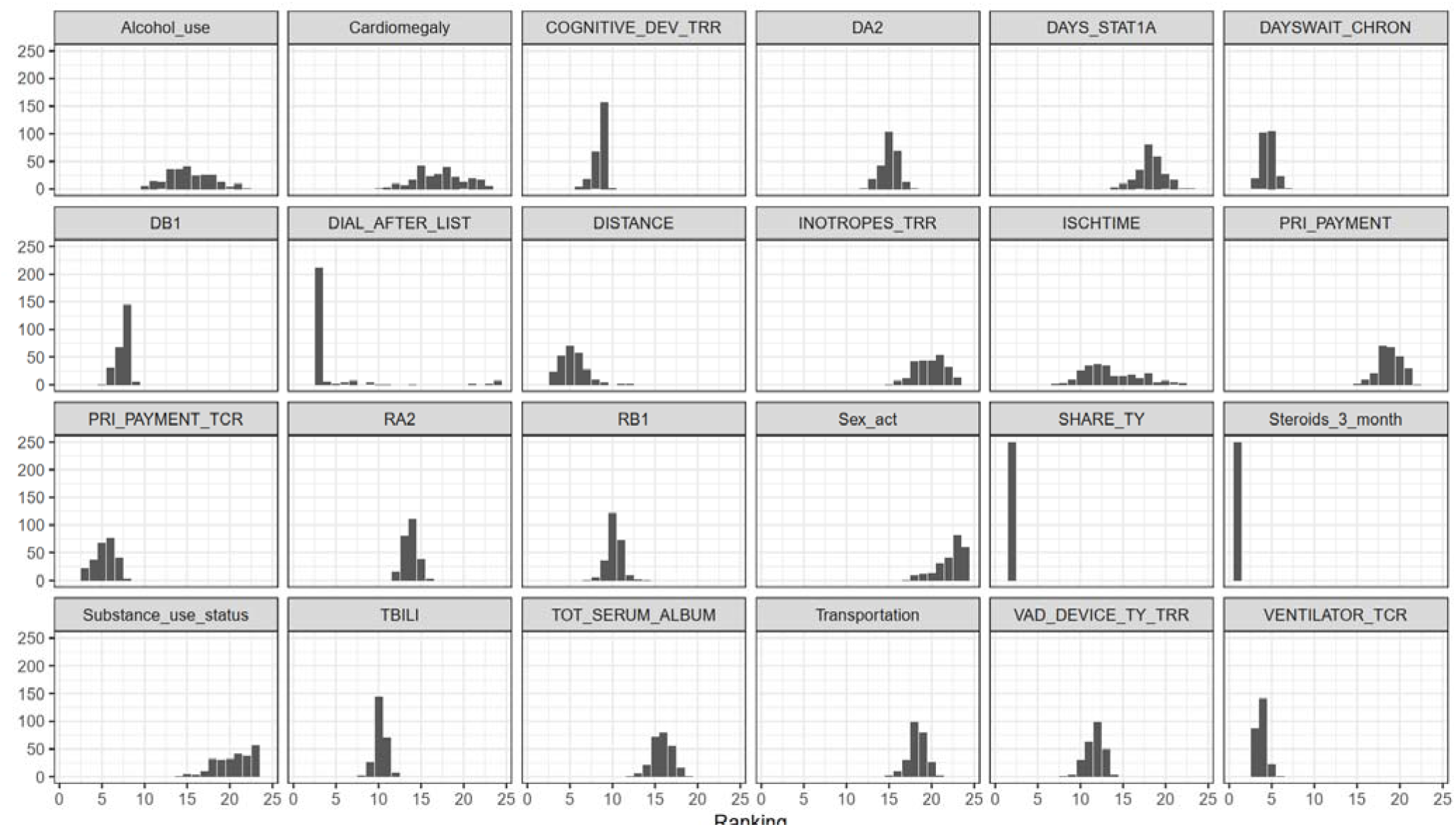
Frequency of ranking of each variable for the 3-year rejection based on pairwise comparison of model reliance. Variables are arranged by average ShapleyVIC values.

**Figure 9.**
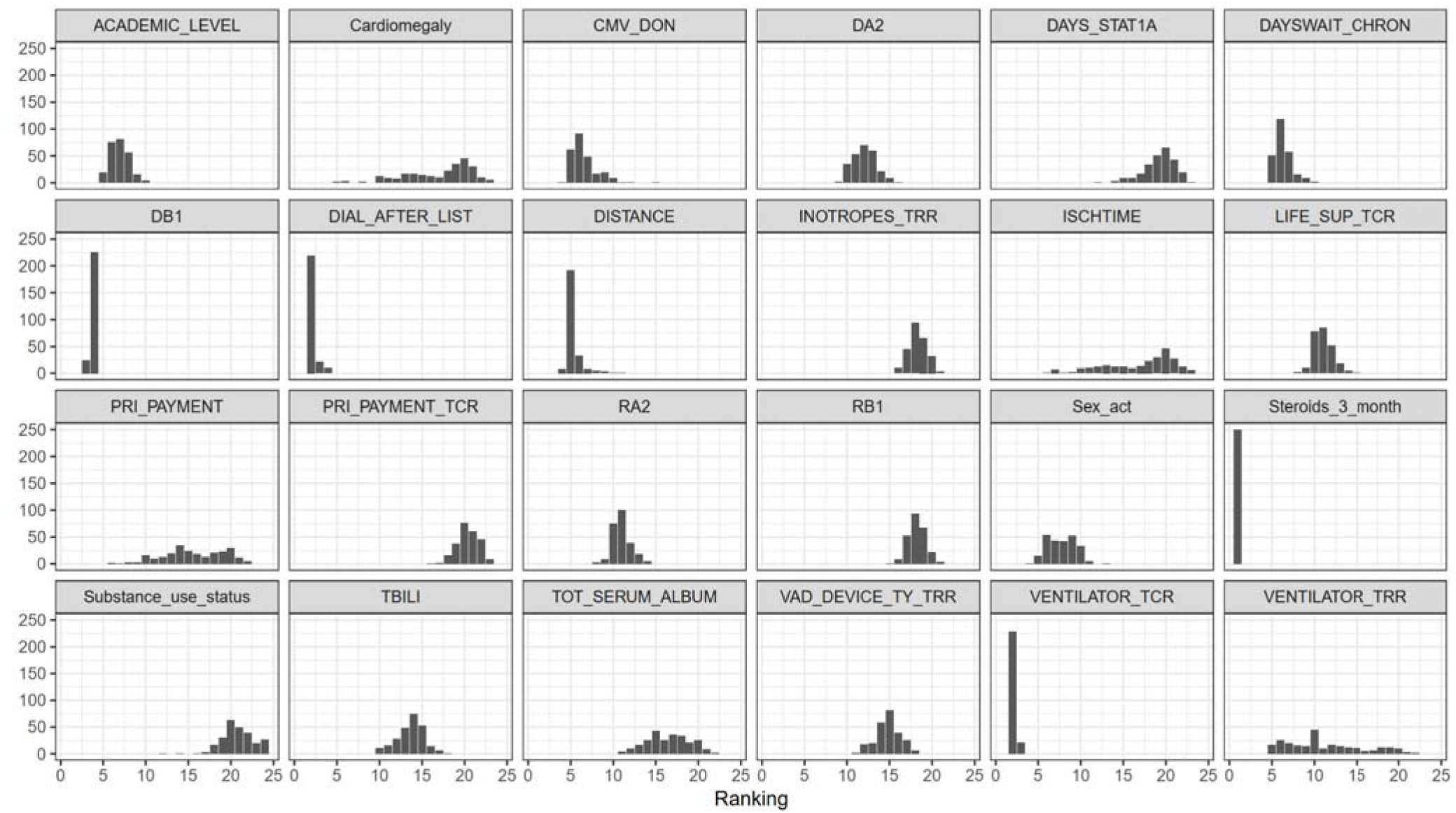
Frequency of ranking of each variable for the 5-year rejection based on pairwise comparison of model reliance. Variables are arranged by average ShapleyVIC values.

We conducted a final model evaluation using a logistic regression model with the selected 20 features to predict organ rejection at 1-, 3-, and 5-year intervals post-transplant. The evaluation results, presented in Table 6, demonstrate that the model exhibits ideal calibration, as evidenced by a calibration intercept of 0.000, indicating no overall bias in predictions, and a calibration slope of 1.000, confirming a perfect proportional relationship between predicted probabilities and actual risks. Furthermore, the model’s C-statistic (AUC) values, ranging from 0.766 to 0.815, reflect moderate to high predictive performance, suggesting reliable discrimination between patients who will experience rejection and those who will not.

**Table 6:** Calibration and Discrimination Metrics for the Logistic Regression Model Using the Selected 20 Features.

|  | Calibration Intercept | Calibration Slop | C-stat (AUC) |
| --- | --- | --- | --- |
| 1-year rejection | 0.000 | 1.000 | 0.766 |
| 3-year rejection | 0.000 | 1.000 | 0.815 |
| 5-year rejection | 0.000 | 1.000 | 0.814 |

## Discussion

Pediatric heart transplantation faces persistent challenges of limited donor organ availability, prolonged waitlist times, and high risks of LAR and hospitalization[4,29–31]. While existing predictive models primarily rely on registry data, their limited inclusion of evolving clinical parameters and social factors restricts their accuracy and utility[17,32,33]. This study addresses these gaps by integrating EHR and SDoH with UNOS registry data, enabling a more holistic and dynamic approach to predicting post-transplant outcomes.

This study shed light on factors influencing pediatric heart transplant rejection, focusing on the interconnected clinical, social, and behavioral aspects that impact patient outcomes. The overall pattern we observe in 1,3- and 5-year models is that the use of life support, like mechanical ventilation, often signals severe illness and multisystem dysfunction, which complicates care post-transplant. Patients requiring life support are typically in critical condition and may need more intensive monitoring and management post-transplant to mitigate rejection risk and enhance outcomes [34]. Longer waiting times on the transplant list are associated with a higher rejection risk due to the deterioration of the patient’s health [35,36], which may lead to increased hospitalizations and interventions like mechanical circulatory support. These factors can increase anti-human leukocyte antigen (HLA) antibody production, which complicates post-transplant outcomes. This is further compounded by the stress and resource limitations faced by patients and caregivers, elevating the overall burden and rejection risk [33,37]. Cognitive development significantly impacts a patient’s or parents/caregiver’s ability to manage chronic conditions and adhere to complex post-transplant regimens. Impaired cognitive function and substance use, for example, can lead to high-risk behaviors such as medication non-adherence or missed follow-ups. In pediatric cases, these behaviors increase the likelihood of post-transplant complications and rejection, emphasizing the need for early psychological and behavioral interventions [32,37].

We also found that patients who spend extended time at Status One (a high-priority transplant status) generally have more aggressive pre-transplant interventions, such as transfusions and desensitization therapies, increasing the likelihood of donor-specific antibody development. While these treatments are crucial for survival, they elevate immunologic risks, making post-transplant rejection more probable [34]. Lab tests predictive features revealed that elevated serum bilirubin and creatinine levels signal underlying organ dysfunction, like liver or kidney issues, which may arise from systemic illnesses or chronic heart failure. These markers indicate compromised overall health, which increases the risk of post-transplant complications. Monitoring these and other organ function markers can help predict and proactively address post-transplant risks.

We also see a strong trend of social determinants, demographics, and behavioral features coming as important predictors. Factors such as transportation challenges, alcohol use, and insurance type—alongside health-related measures like BMI—play significant roles in post-transplant outcomes. Higher BMI is linked to metabolic complications [38], while transportation issues can lead to missed follow-ups [39], negatively impacting patient adherence to post-transplant care. Insurance type serves as a proxy for socioeconomic status, with Medicaid often reflecting limited healthcare access [33,40]. Social challenges like these can lead to suboptimal care, necessitating targeted interventions to address disparities in access and support for pediatric transplant patients. Insurance coming as a strong predictor in 3 and 5 years shows that the patient received good treatment in the initial time of transplant but due to not having good insurance may affect further follow-up or medication adherence in the following 3-5 years. The findings underscore the need for a holistic approach to managing pediatric transplant patients, considering both medical and social factors that contribute to rejection risk [41,42]. Clinical strategies should integrate psychological support, social services, and tailored monitoring for high-risk patients, such as those with prolonged waiting times or those needing life support pre-transplant. Addressing socioeconomic disparities and enhancing access to comprehensive care—through targeted support for transportation, adherence programs, and access to preventative care—are essential for improving long-term transplant outcomes.

## Limitations and Future Enhancements

This study has several limitations that warrant consideration. First, the use of a single-center dataset limits the generalizability of the findings to other pediatric transplant populations, necessitating validation across multicenter cohorts. Second, the reliance on proxy measures for social determinants of health (SDoH), such as insurance type, may not fully capture the nuanced socioeconomic and behavioral factors influencing outcomes. Lastly, the study spans nearly three decades, introducing potential variability due to changes in medical practices and transplantation policies. Addressing these limitations in future research will enhance the robustness and applicability of the findings.

## Conclusions

In conclusion, this study highlights the multifaceted clinical, social, and behavioral factors influencing pediatric heart transplant rejection and underscores the importance of a holistic approach to post-transplant care. Machine learning models integrating electronic health records (EHR), social determinants of health (SDoH), and United Network for Organ Sharing (UNOS) data demonstrated superior predictive performance compared to models using registry data alone, achieving weighted F1-scores of 0.881, 0.835, and 0.884 for predicting rejection at 1-, 3-, and 5-year intervals, respectively. Key findings reveal severe pre-transplant conditions, such as the use of life support, prolonged waitlist times, and aggressive interventions, elevate rejection risk due to multisystem dysfunction and immunologic complications. Additionally, elevated bilirubin and creatinine levels indicate underlying organ dysfunction and heightened vulnerability to post-transplant complications. Social and behavioral factors, including transportation challenges, BMI, and insurance type, were also critical predictors, reflecting disparities in healthcare access and adherence to care regimens. Notably, Medicaid coverage emerged as a significant factor in long-term rejection outcomes, highlighting the influence of socioeconomic status on sustained post-transplant care. The findings emphasize the need for integrated care strategies that address medical and social health determinants. Tailored monitoring, psychological and behavioral support, and targeted interventions to mitigate disparities in access and adherence are crucial for improving long-term outcomes. This study underscores the potential of data-driven, multidisciplinary approaches to enhance personalized care and reduce the burden of transplant rejection in pediatric patients.

## Data Availability

Due to the HIPPA regulation, the study data cannot be shared.

## Acknowledgements

This project was partially supported by the National Library of Medicine under grant number R21LM013911, and the University of Florida-Florida State University Clinical and Translational Science Award, which is supported in part by the NIH National Center for Advancing Translational Sciences under award number UL1TR001427. The content is solely the responsibility of the authors and does not necessarily represent the official views of the National Institutes of Health.

